# The Association Between Social Determinants of Health and Alzheimer Disease Blood Biomarkers in Midlife

**DOI:** 10.64898/2026.04.13.26350798

**Authors:** Christina S. Dintica, Gargi Porwal, Michelle Caunca, Nathaniel Fleming, R. Nick Bryan, Kristine Yaffe

**Affiliations:** Department of Psychiatry, University of California, San Francisco, San Francisco, CA, United States; Department of Neurology, Weill Institute for Neurosciences, University of California San Francisco, San Francisco, CA, USA; Department of Radiology, University of Pennsylvania, Philadelphia, PA, USA; Department of Epidemiology and Biostatistics, University of California San Francisco

## Abstract

**Background:** Social determinants of health (SDOH) are increasingly recognized as contributors to Alzheimer disease (AD) risk, yet the impact of multidimensional social disadvantage early AD-related pathophysiology remains poorly understood.

**Methods:** We studied 1,466 participants from the Coronary Artery Risk Development in Young Adults (CARDIA) cohort with SDOH assessed in early midlife (mean age 40 ± 3.6 years) and plasma AD biomarkers measured 20 years later. A comprehensive SDOH index was constructed from 12 indicators spanning five domains (economic stability, education, neighborhood and physical environment, community and social context, and health care access). We examined associations between SDOH quartile and log-transformed, standardized plasma phosphorylated tau 217 (p-tau217), neurofilament light chain (NfL), and amyloid-β42/40 (Aβ42/40) using linear regression adjusted for age, sex, race, and estimated glomerular filtration rate. Linear trends across SDOH quartile were also evaluated.

**Results:** Participants in the most disadvantaged SDOH quartile had higher p-tau217, higher NfL and lower Aβ42/40 level compared with those in the least disadvantaged quartile (p-tau 217: β = 0.12, 95% CI 0.03–0.21, p = 0.008; NfL: β = 0.20, 95% CI 0.05–0.35, p = 0.009; Aβ42/40: β = −0.15, 95% CI −0.30-0.00, p=0.05). There was also a significant trend across quartile (p-tau 217: *p* for trend = 0.012; NfL: p for trend =0.001). Analyses of individual SDOH domains indicated that lower economic stability, poorer health care access, and lower education were associated with higher NfL, and poorer health care access was associated with higher p-tau217.

**Conclusions:** Greater SDOH disadvantage in early midlife was associated with higher levels of plasma AD biomarkers reflecting AD pathology and neurodegeneration decades later. These findings suggest that social disadvantage during midlife may contribute to early AD-related biological changes and highlight potentially modifiable social factors relevant for dementia prevention.

## INTRODUCTION

Social determinants of health (SDOH) encompass the economic, social, and environmental conditions that shape health outcomes across the life course. Factors such as socioeconomic status (SES), educational attainment, employment, housing, and access to health care are increasingly recognized as important contributors to Alzheimer disease (AD) risk.^1,2^

Despite this growing body of evidence, studies examining the impact of multiple SDOH domains on AD-related pathology remain limited. Existing composite indices often emphasize economic indicators while underrepresenting social, environmental, and health care–related conditions.^8^ In addition, most prior studies have focused on late-life populations, leaving midlife—a critical but understudied period—relatively unexplored. Neuropathological processes associated with AD are known to begin decades before clinical symptom onset, making midlife a key window for identifying early social risk factors.^9,10^

To address these gaps, we leveraged longitudinal data from the Coronary Artery Risk Development in Young Adults (CARDIA) study to construct a multidimensional SDOH index aligned with the Healthy People 2030 framework, incorporating domains of economic stability, education, neighborhood and physical environment, community and social context, and health care access and quality.^14^ Using this index, we examined whether adverse SDOH in early midlife are associated with plasma AD biomarkers (Aβ, p-tau, NfL) 20 years later. This multidomain approach in a well-characterized cohort highlights potentially modifiable social factors influencing early AD-related changes and informs prevention efforts.

## METHODS

### Study Population

The Coronary Artery Risk Development in Young Adults (CARDIA) study is a prospective cohort study investigating the development of and risk factors for cardiovascular disease.^17^ Briefly, starting in 1985, 5115 Black and White adults between 18 and 30 years of age were recruited from population-based samples of 4 US cities (Birmingham, AL; Chicago, IL; Minneapolis, MN; and Oakland, CA). Within each center, recruitment was balanced by sex, age, and educational level. Participants completed follow-up exams every 2–5 years over 30 years (Years 2–35; 1987–2022).

At each examination, participants provided written informed consent, and study protocols were approved by institutional review boards at each study site and the CARDIA Coordinating Center. Further details regarding the design and recruitment of CARDIA have been previously reported. For this study, we included 1,466 participants from CARDIA who had SDOH data assessed during early midlife and plasma AD biomarkers measured 20 years later.

### SDOH index

As a summary of social burden, we created an aggregate SDOH index based on the framework described by the Healthy People 2030 initiative, which categorizes SDOH into 5 domains: economic stability, neighborhood and physical environment, education, community and social context, and health care access and quality.^14^ In CARDIA, SDOH domains included economic stability (income, housing, financial strain), education (years), community/social context (support, stress, cohesion, discrimination), health care access (care source, insurance, costs), and neighborhood resources (activity access). Twelve items were dichotomized (1 = adverse, 0 = not) and weighted so each domain contributed equally (0–0.2) (Supplementary Table 1). Domain scores were summed to create an index (0–1), with higher values indicating greater disadvantage, and categorized into quartiles (Q1 least, Q4 most disadvantaged). SDOH measures were assessed at years 15/20 (averaged if both available).

### Blood Processing and Plasma Biomarkers

Blood samples were drawn at the Year 35 clinic visit, processed and stored within 90 min (stick-to-freezer) at -70°C until assayed between March and May 2024 at the Laboratory of Pathology and Laboratory Medicine at the Hospital of the University of Pennsylvania. Plasma Aβ1-42, Aβ1-40, and p-tau217 were quantified using the Fujirebio fully automated Lumipulse chemiluminescence enzyme immunoassay platform (Lumipulse G1200, Fujirebio Diagnostics) with results reported in pg/mL within the manufacturer’s specified ranges.

To ensure quality assurance, calibration standards and manufacturer-provided aqueous quality controls (QCs) were included in each analytical run. In addition, six plasma pool QC samples derived from individuals with a range of biomarker profiles were analyzed in parallel. Across 15 analytical runs, the average concentrations and coefficients of variation (%CV) for p-tau217, Aβ1-42, and Aβ1-40 were consistently within acceptable limits (e.g., Con1: p-tau217 = 0.452 pg/mL, %CV = 3.76; Aβ1-42 = 20.34 pg/mL, %CV = 3.84; Aβ1-40 = 212.88 pg/mL, %CV = 2.87). Full QC performance data are summarized in Supplementary Table 2.

### Covariates

Demographic characteristics and cigarette smoking (current or former) were based on self-report. Body mass index (BMI) was calculated as weight in kilograms divided by height in meters squared. Diabetes mellitus at baseline was defined as fasting plasma glucose ≥126 mg/dL, oral glucose tolerance test ≥200 mg/dL, glycosylated hemoglobin ≥6.5%, or use of diabetes medications. A digital blood pressure monitor (Omron HEM-907XL; Online Fitness) was used to measure systolic and diastolic blood pressure. Hypertension was defined as systolic BP ≥130 mmHg, diastolic BP ≥80 mmHg. Estimated glomerular filtration rate (eGFR) was calculated using the 2021 CKD-EPI creatinine equation without race, which incorporates serum creatinine, age, and sex to estimate kidney function in mL/min/1.73 m^2.19^

### Statistical Analysis

Participant characteristics across SDOH quartile were compared using χ^2^ tests for categorical variables and one-way analysis of variance (ANOVA) for continuous variables. For biomarker analyses, linear trends across SDOH quartile were assessed by modeling the ordinal quartile variable as a continuous term and reporting the corresponding *p*-value for trend.

Blood-based biomarkers were log-transformed to address skewed distributions and subsequently standardized (z-scored) to facilitate comparison of effect sizes across biomarkers. Associations between SDOH quartile and biomarkers were examined using linear regression models with robust standard errors. Model-adjusted marginal means and 95% confidence intervals were derived using post-estimation margins.

Primary models were adjusted for age, sex, race, and estimated glomerular filtration rate (eGFR), given prior evidence linking kidney function to circulating AD biomarker levels. Additional models further adjusted for vascular risk factors (including hypertension, diabetes, smoking status, body mass index) and depressive symptoms.

Effect modification by sex and race was assessed by including interaction terms between the SDOH index and sex and race, respectively, in linear regression models adjusted for age and eGFR.

Separate linear regression models were used to evaluate associations between individual SDOH domains, modeled as continuous standardized variables, and each biomarker. These models were adjusted for age, sex, race, and eGFR.

All statistical analyses were conducted using Stata version 15.1 (StataCorp, College Station, TX) and RStudio version 4.0.4 (R Foundation for Statistical Computing, Vienna, Austria).

## RESULTS

Participants had a mean age of 40 years (range 32–52); 57% (n = 833) were female and 44% (n = 646) identified as Black. The SDOH index was slightly left-skewed, with a median of 0.17 (interquartile range: −0.10 to 0.59). Participants in higher SDOH disadvantage quartile were more likely to be younger, female, and Black, and exhibited a less favorable cardiometabolic and behavioral risk profile, including higher prevalence of smoking, hypertension, diabetes, higher BMI, and lower levels of physical activity (Table 1).

**Table 1.** Study Population Characteristics by SDOH Quartile (N = 1,466).

| Characteristic | Q1<br>least disadvantage<br>(n=463) | Q2<br>(n=348) | Q3<br>(n=328) | Q4<br>Most disadvantaged<br>(n=327) | p-value |
| --- | --- | --- | --- | --- | --- |
| Age, Mean (SD) | 40.98 (3.38) | 40.30 (3.64) | 40.36 (3.68) | 39.93 (3.78) | 0.001 |
| Female, n (%) | 237 (51.2) | 211 (60.6) | 195 (59.5) | 190 (58.1) | 0.027 |
| Black race, n (%) | 105 (22.7) | 152 (43.7) | 179 (54.6) | 210 (64.2) | <0.001 |
| Diabetes, n (%) | 14 (3.1) | 12 (3.5) | 24 (7.7) | 34 (11.1) | <0.001 |
| Smoking, n (%) | 130 (28.1) | 116 (33.3) | 130 (39.8) | 166 (50.8) | <0.001 |
| Hypertension | 26 (5.6) | 40 (11.5) | 29 (8.8) | 43 (12.8) | 0.002 |
| BMI, Mean (SD) | 27.58 (6.28) | 28.43 (6.12) | 29.62 (6.90) | 30.03 (7.27) | <0.001 |
| Apolipoprotein e4 | 122 (28.8) | 99 (31.0) | 100 (34.0) | 98 (33.0) | 0.460 |

Participants in the highest SDOH disadvantage quartile exhibited higher plasma p-tau217 levels compared with those in the lowest quartile (β = 0.12, 95% CI 0.03–0.21, *p* = 0.008), There was evidence of an overall linear trend across SDOH quartile for p-tau217 (*p* for trend = 0.012). A similar pattern was observed for neurofilament light chain (NfL), with higher levels among participants in the most disadvantaged quartile relative to the least disadvantaged (β = 0.20, 95% CI 0.05–0.35, *p* = 0.009), and a significant trend across quartile (*p* for trend = 0.001).

Participants in the highest disadvantage quartile also tended to have a lower Aβ42/40 ratio (β = −0.15, 95% CI −0.30 to 0.00, *p* = 0.052, (*p* for trend across quartile= 0.064). Adjusted means are shown in Table 2. Additional adjustment for vascular risk factors slightly attenuated the results but remained similar (Table 2).

**Table 2.**
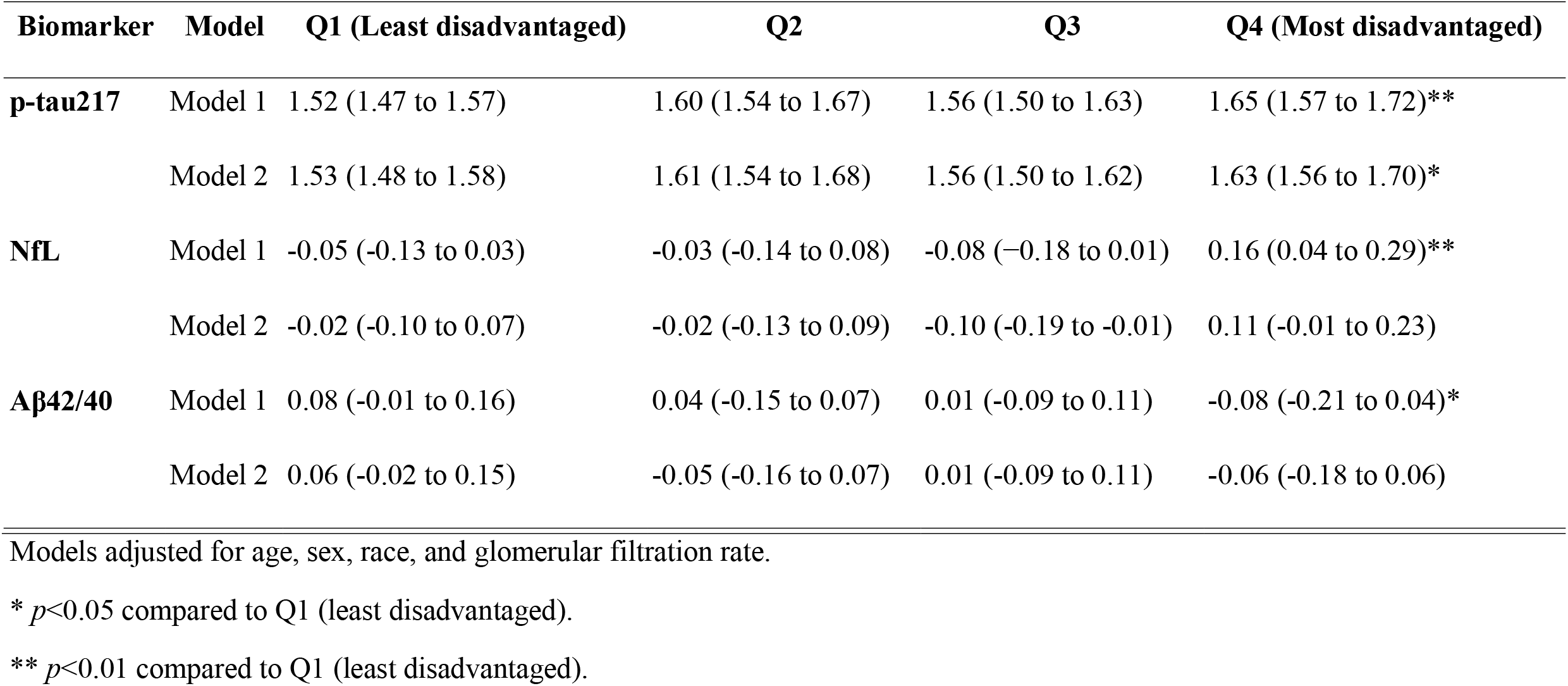
Adjusted mean levels of AD blood biomarkers across SDOH quartile (95% confidence intervals).

| **Biomarker** | **Model** | **Q1 (Least disadvantaged)** | **Q2** | **Q3** | **Q4 (Most disadvantaged)** |
| --- | --- | --- | --- | --- | --- |
| **p-tau217** | Model 1 | 1.52 (1.47 to 1.57) | 1.60 (1.54 to 1.67) | 1.56 (1.50 to 1.63) | 1.65 (1.57 to 1.72)** |
|  | Model 2 | 1.53 (1.48 to 1.58) | 1.61 (1.54 to 1.68) | 1.56 (1.50 to 1.62) | 1.63 (1.56 to 1.70)* |
| **NfL** | Model 1 | -0.05 (-0.13 to 0.03) | -0.03 (-0.14 to 0.08) | -0.08 (-0.18 to 0.01) | 0.16 (0.04 to 0.29)** |
|  | Model 2 | -0.02 (-0.10 to 0.07) | -0.02 (-0.13 to 0.09) | -0.10 (-0.19 to -0.01) | 0.11 (-0.01 to 0.23) |
| **Aβ42/40** | Model 1 | 0.08 (-0.01 to 0.16) | 0.04 (-0.15 to 0.07) | 0.01 (-0.09 to 0.11) | -0.08 (-0.21 to 0.04)* |
|  | Model 2 | 0.06 (-0.02 to 0.15) | -0.05 (-0.16 to 0.07) | 0.01 (-0.09 to 0.11) | -0.06 (-0.18 to 0.06) |

Models adjusted for age, sex, race, and glomerular filtration rate.
\* $p < 0.05$ compared to Q1 (least disadvantaged).
\*\* $p < 0.01$ compared to Q1 (least disadvantaged).

In analyses of individual SDOH domains, we found that less economic stability (β: 0.10, 95% CI: 0.04 to 0.16), less healthcare access (β: 0.10, 95% CI: 0.04 to 0.15), and less education (β: 0.09, 95% CI: 0.02 to 0.15) were independently associated with higher NfL. Less healthcare access was also associated with higher ptau-217 (β: 0.04, 95% CI: 0.01 to 0.08).

No interactions were observed between the SDOH index and sex or race in relation to AD biomarkers (all p > 0.10).

## DISCUSSION

In this study of community-dwelling adults, greater social disadvantage in early midlife was associated with higher levels of AD–related plasma biomarkers measured approximately 20 years later. Participants in the highest SDOH disadvantage quartile had higher levels of p-tau217 and neurofilament light chain (NfL), with a similar but weaker pattern observed for Aβ42/40. Together, these findings suggest that social disadvantage may be associated with early biological changes relevant to both neurodegeneration and AD pathology, detectable before the onset of clinical symptoms.^9^

The pattern of associations across biomarkers—strongest for NfL, moderate for p-tau217, and weaker for Aβ42/40—is consistent with current models of AD progression.^9^ In midlife, variability in amyloid burden is often limited, and age remains a dominant determinant of amyloid accumulation,^20,21^ which may reduce power to detect associations with Aβ42/40. In contrast, markers of neurodegeneration and neuronal injury, such as NfL, may be more sensitive to exposures during this period. NfL is a well-established marker of neuroaxonal injury across neurological conditions,^22,23^ and has been linked to structural and vascular-related brain changes.^24^ These findings suggest that social disadvantage may be more strongly reflected in downstream neurodegenerative processes than in early amyloid accumulation in midlife populations.

Our study is among the first to examine a comprehensive, multidomain SDOH index spanning all five Healthy People 2030 domains^14^ in relation to AD-related biomarkers in midlife. Analyses of individual SDOH domains showed that economic stability, health care access, and education are particularly relevant, however, no single domain explained the associations, highlighting the importance of considering multidomain social disadvantage rather than isolated factors.

Strengths of this study include its population-based design, large sample of adults in midlife, and the use of a multidomain SDOH measure. Several limitations should be noted. Residual confounding by unmeasured life-course factors, including early-life socioeconomic conditions and wealth, is possible. Selection and attrition over long-term follow-up may have influenced the analytic sample. Biomarkers were measured at a single time point, limiting assessment of trajectories. In addition, while vascular risk factors may play an important role, their dual role as confounders and potential mediators complicates interpretation. Finally, the cohort includes only Black and White participants, which may limit generalizability to other populations.

In summary, social disadvantage in early midlife was associated with modestly higher levels of p-tau217 and NfL measured approximately two decades later. These findings suggest that social disadvantage may be biologically embedded by midlife and reflected in AD-related and neurodegenerative biomarker profiles well before clinical symptoms emerge. Addressing social determinants of health earlier in the life course may therefore represent an important strategy for reducing disparities in dementia risk.

**Figure 1.**
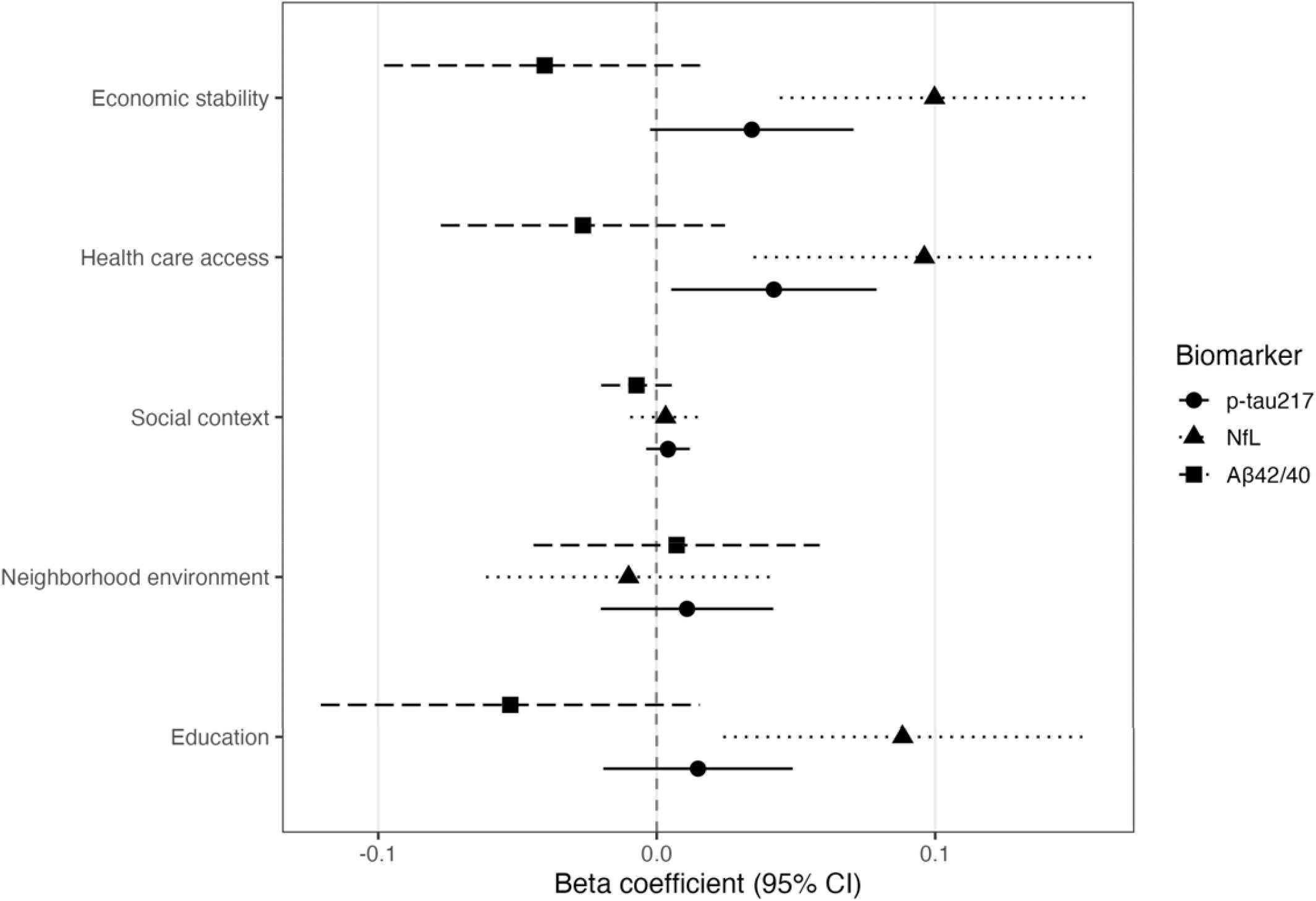
Association between individual SDOH domains and AD blood biomarkers. Models adjusted for age, sex, race, and glomerular filtration rate.

## Data Availability

The CARDIA data can be requested via the study website https://www.cardia.dopm.uab.edu/. Data access is available through the CARDIA Coordinating Center following approval by the CARDIA Publications and Presentations Committee. Please see the study website for further details: http://www.cardia.dopm.uab.edu/invitation-to-new-investigators.

## ACKNOWLEDGEMENTS

The Coronary Artery Risk Development in Young Adults Study (CARDIA) is supported by contracts 75N92023D00002, 75N92023D00003, 75N92023D00004, 75N92023D00005, and 75N92023D00006 from the National Heart, Lung, and Blood Institute (NHLBI).

CARDIA was also partially supported by the Intramural Research Program of the National Institute on Aging (NIA) and an intra-agency agreement between NIA and NHLBI (AG0005).

## REFERENCES

1. Walsh S, Klee M, Hui EK, et al. Social determinants of dementia: A scoping review. Alzheimers Dement. 2025;21(7):e70524. doi:10.1002/alz.70524

2. Wang K, Fang Y, Zheng R, et al. Associations of socioeconomic status and healthy lifestyle with incident dementia and cognitive decline: two prospective cohort studies. eClinicalMedicine. 2024;76:102831. doi:10.1016/j.eclinm.2024.102831

3. Myoraku A, Hausle I, Mila-Aloma M, et al. Association of neighborhood disadvantage with Alzheimer’s disease pathology and the stability of blood-based biomarker performance. J Prev Alzheimers Dis. 2026;13(2):100445. doi:10.1016/j.tjpad.2025.100445

4. Marks SM. Association of Lifetime Cognitive Engagement and Low β-Amyloid Deposition. Arch Neurol. 2012;69(5):623. doi:10.1001/archneurol.2011.2748

5. Powell WR, Zuelsdorff M, Keller SA, et al. Association of Neighborhood-Level Disadvantage With Neurofibrillary Tangles on Neuropathological Tissue Assessment. JAMA Netw Open. 2022;5(4):e228966. doi:10.1001/jamanetworkopen.2022.8966

6. Shen C, Rolls ET, Cheng W, et al. Associations of Social Isolation and Loneliness With Later Dementia. Neurology. 2022;99(2). doi:10.1212/WNL.0000000000200583

7. Choi EY, Cho G, Chang VW. Neighborhood Social Environment and Dementia: The Mediating Role of Social Isolation. Kelley JA, ed. J Gerontol B Psychol Sci Soc Sci. 2024;79(4):gbad199. doi:10.1093/geronb/gbad199

8. Morenz AM, Liao JM, Au DH, Hayes SA. Area-Level Socioeconomic Disadvantage and Health Care Spending: A Systematic Review. JAMA Netw Open. 2024;7(2):e2356121. doi:10.1001/jamanetworkopen.2023.56121

9. Jack CR, Bennett DA, Blennow K, et al. NIALJAA Research Framework: Toward a biological definition of Alzheimer’s disease. Alzheimers Dement. 2018;14(4):535–562. doi:10.1016/j.jalz.2018.02.018

10. Yaffe K, Vittinghoff E, Pletcher MJ, et al. Early Adult to Midlife Cardiovascular Risk Factors and Cognitive Function. Circulation. 2014;129(15):1560–1567. doi:10.1161/CIRCULATIONAHA.113.004798

11. Ren Y, Savadlou A, Park S, Siska P, Epp JR, Sargin D. The impact of loneliness and social isolation on the development of cognitive decline and Alzheimer’s Disease. Front Neuroendocrinol. 2023;69:101061. doi:10.1016/j.yfrne.2023.101061

12. Palpatzis E, Akinci M, AguilarLJDominguez P, et al. Lifetime Stressful Events Associated with Alzheimer’s Pathologies, Neuroinflammation and Brain Structure in a Risk Enriched Cohort. Ann Neurol. 2024;95(6):1058–1068. doi:10.1002/ana.26881

13. Gogniat MA, Khan OA, Ratangee B, et al. Cross-Sectional and Longitudinal Associations of Neighborhood Disadvantage With Fluid Biomarkers of Neuroinflammation and Neurodegeneration. Neurology. 2025;105(2):e213770. doi:10.1212/WNL.0000000000213770

14. U.S. Department of Health and Human Services, Office of Disease Prevention and Health Promotion. Healthy People 2030. https://odphp.health.gov/healthypeople/objectives-and-data/social-determinants-health

15. Gottesman RF, Wu A, Coresh J, et al. Associations of Vascular Risk and Amyloid Burden with Subsequent Dementia. Ann Neurol. 2022;92(4):607–619. doi:10.1002/ana.26447

16. Rabin JS, Yang H, Schultz AP, et al. Vascular Risk and β LJAmyloid Are Synergistically Associated with Cortical Tau. Ann Neurol. 2019;85(2):272–279. doi:10.1002/ana.25399

17. Friedman GD, Cutter GR, Donahue RP, et al. Cardia: study design, recruitment, and some characteristics of the examined subjects. J Clin Epidemiol. 1988;41(11):1105–1116. doi:10.1016/0895-4356(88)90080-7

18. Reis JP, Loria CM, Launer LJ, et al. Cardiovascular health through young adulthood and cognitive functioning in midlife. Ann Neurol. 2013;73(2):170–179. doi:10.1002/ana.23836

19. Inker LA, Eneanya ND, Coresh J, et al. New Creatinine- and Cystatin C–Based Equations to Estimate GFR without Race. N Engl J Med. 2021;385(19):1737–1749. doi:10.1056/NEJMoa2102953

20. Rippon B, Palta P, Tahmi M, et al. Plasma Amyloid and in vivo Brain Amyloid in Late Middle-Aged Hispanics. J Alzheimers Dis. 2022;87(3):1229–1238. doi:10.3233/JAD-210391

21. Gonzales MM, O’Donnell A, Ghosh S, et al. Associations of cerebral amyloid beta and tau with cognition from midlife. Alzheimers Dement. 2024;20(9):5901–5911. doi:10.1002/alz.14060

22. Gaetani L, Blennow K, Calabresi P, Di Filippo M, Parnetti L, Zetterberg H. Neurofilament light chain as a biomarker in neurological disorders. J Neurol Neurosurg Psychiatry. 2019;90(8):870–881. doi:10.1136/jnnp-2018-320106

23. Turner MR, Thompson AG, Teunissen CE. Blood level of neurofilament light chain as a biomarker for neurological disorders. BMJ Med. 2025;4(1):e000958. doi:10.1136/bmjmed-2024-000958

24. Gisslén M, Price RW, Andreasson U, et al. Plasma Concentration of the Neurofilament Light Protein (NFL) is a Biomarker of CNS Injury in HIV Infection: A Cross-Sectional Study. EBioMedicine. 2016;3:135–140. doi:10.1016/j.ebiom.2015.11.036

25. Lewis TT, Cogburn CD, Williams DR. Self-Reported Experiences of Discrimination and Health: Scientific Advances, Ongoing Controversies, and Emerging Issues. Annu Rev Clin Psychol. 2015;11(1):407–440. doi:10.1146/annurev-clinpsy-032814-112728

26. McEwen BS. Neurobiological and Systemic Effects of Chronic Stress. Chronic Stress. 2017;1:2470547017692328. doi:10.1177/2470547017692328

27. González HM, Tarraf W, Harrison K, et al. Midlife cardiovascular health and 20LJyear cognitive decline: Atherosclerosis Risk in Communities Study results. Alzheimers Dement. 2018;14(5):579–589. doi:10.1016/j.jalz.2017.11.002

28. Kind AJH, Buckingham WR. Making Neighborhood-Disadvantage Metrics Accessible — The Neighborhood Atlas. N Engl J Med. 2018;378(26):2456–2458. doi:10.1056/NEJMp1802313

29. Flanagan BE, Gregory EW, Hallisey EJ, Heitgerd JL, Lewis B. A Social Vulnerability Index for Disaster Management. J Homel Secur Emerg Manag. 2011;8(1):0000102202154773551792. doi:10.2202/1547-7355.1792

30. Desai P, Ng TKS, Krueger KR, Evans DA, Rajan KB. Social Vulnerability Index and Cognitive Decline in a PopulationLJBased Cohort Study. Alzheimers Dement. 2024;20(S7):e089070. doi:10.1002/alz.089070

