## Supplementary Table 1 for "The Association Between Social Determinants of Health and Alzheimer Disease Blood Biomarkers in Midlife"

**Supplementary Table 1.** Itemized questionnaire for social determinants of health domains in CARDIA.

| **Domain** | **Individual item** | **Survey response/scale** | **Analytic recode** |
| --- | --- | --- | --- |
| **ECONOMIC STABILITY** | Household Income: Gross family income, past 12 months | <$5,000; $5,000-$7,999; $8,000-$11,999; $12,000-$15,999; 16000-19999; 20000-24,999; 25000-29999; 30000-34999; 35000-39999; 40000-49999; 50000-74999; 75000-99999; 100000+ | 0=≥$50,000  1=<$50,000 |
|  | House Tenure: Residence-Own or rent | Rent; Pay a mortgage; Own free and clear; Other No | 0 = "Pay a mortgage/Own free and clear"; 1 = "Rent/other" |
|  | Financial strain: Ongoing financial strain | Yes; No | 0 = "No"; 1 = "Yes" |
| **EDUCATION** | Education: Highest level completed | Elementary school: Grades 1-8; Highschool: grades 9-10; College: 13-16; Graduate school: 17-20+ | 0=“≥Some college” 1=“≤High school” |
| **COMMUNITY & SOCIAL SUPPORT** |  |  |  |
|  | Social Support: Measured by the Emotional Social Support Index (From the following 5 items) |  | From the aggregate sum of the following 9 items, divided into tertiles: 0”high emotional/social support”= tertile 0-1; 1= “Low emotional/social support” |
|  | Someone available to listen to you | None of the time; A little of the time; Some of the time; Most of the time; All of the time | 1= “None of the time”; 2= “A little of the time”; 3= “Some of the time”; 4=“Most of the time”; 5= “All of the time” |
|  | Someone available to give you advice | None of the time; A little of the time; Some of the time; Most of the time; All of the time | 1= “None of the time”; 2= “A little of the time”; 3= “Some of the time”; 4= “Most of the time”; 5= “All of the time” |
|  | Someone available to show you love and affection | None of the time; A little of the time; Some of the time; Most of the time; All of the time | 1= “None of the time”; 2= “A little of the time”; 3= “Some of the time”; 4=“Most of the time”; 5= “All of the time” |
|  | Someone available to give you emotional support | None of the time; A little of the time; Some of the time; Most of the time; All of the time | 1= “None of the time”; 2= “A little of the time”; 3= “Some of the time”; 4= “Most of the time”; 5= “All of the time” |
|  | Sufficient contact with someone you can confide in | None of the time; A little of the time; Some of the time; Most of the time; All of the time | 1= “None of the time”; 2= “A little of the time”; 3= “Some of the time”; 4= “Most of the time”; 5= “All of the time” |
|  | Chronic stress:  (Assessed using the chronic burden scale).51,52 If participant answered “yes”, they were then asked how severe. Aggregate score of questions in which moderate to severe stress was reported |  | 0= Low/Medium chronic stress; 1= High chronic stress |
|  |  | Ongoing health problems (self), >6 months: Yes; No | 0 = "No"; 1 = "Yes" |
|  |  | Ongoing health problem (someone close to you), > 6 months: Yes; No | 0 = "No"; 1 = "Yes" |
|  |  | Ongoing job difficulties, > 6 months: Yes; No | 0 = "No"; 1 = "Yes" |
|  |  | Ongoing financial strain, > 6 months: Yes; No | 0 = "No"; 1 = "Yes" |
|  |  | Ongoing relationship problems, > 6 months: Yes; No | 0 = "No"; 1 = "Yes" |
|  |  | If “yes” to any of the above: How stressful?: Yes; No | 1= “Not very stressful”; 2= “Moderately stressful”; 3= “Very stressful” |
|  | Discrimination based on gender, race, or socioeconomic status |  | 1= responded yes to any of the questions on discrimination; 0= answered no to all questions on discrimination |
|  | Discrimination: Gender  Have you ever experienced discrimination, been prevented from doing something, or been hassled or made to feel inferior in any of the following seven situations because of your gender? - At school  - Getting a job  - Getting housing  - At home  - Getting medical care  - On the street or in a public setting | Yes; No | 0 = "No"; 1 = "Yes" |
|  | Discrimination: Race  Have you ever experienced discrimination, been prevented from doing something, or been hassled or made to feel inferior in any of the following seven situations because of your race? - At school  - Getting a job  - Getting housing  - At home  - Getting medical care  - On the street or in a public setting | Yes; No | 0 = "No"; 1 = "Yes" |
|  | Discrimination: Socioeconomic status  Have you ever experienced discrimination, been prevented from doing something, or been hassled or made to feel inferior in any of the following seven situations because of your socioeconomic status? - At school  - Getting a job  - Getting housing  - At home  - Getting medical care  - On the street or in a public setting | Yes; No | 0 = "No"; 1 = "Yes" |
|  | Neighborhood Cohesion Composite Score (aggregate score from the following 5 questions): |  | From the aggregate sum of the following 5 items, divided into tertiles: 0 = tertile 1-2; 1= tertile 3 |
|  | Close-knit neighborhood | Strongly agree; Agree; Neither agree nor disagree; Disagree; Strongly disagree | 1=“Strongly agree”;_2=“Agree”; 3=_ “Neither agree nor _disagree”; 4=_ “Disagree”; 5= “Strongly _disagree” _ |
|  | People willing to help their neighbors | Strongly agree; Agree; Neither agree nor disagree; Disagree; Strongly disagree | 1=“Strongly agree”;_2=“Agree”; 3=_ “Neither agree nor _disagree”; 4=_ “Disagree”; 5= “Strongly _disagree” _ |
|  | People in neighborhood don’t get along | Strongly agree; Agree; Neither agree nor disagree; Disagree; Strongly disagree | 1= “Strongly agree”;_2=“Agree”; 3=_ “Neither agree nor _disagree”; 4=_ “Disagree”; 5= “Strongly _disagree” _ |
|  | People in neighborhood can be trusted | Strongly agree; Agree; Neither agree nor disagree; Disagree; Strongly disagree | 1= “Strongly agree”; 2= “Agree”; 3=_ “Neither agree nor _disagree”; 4= “Disagree”; 5= “Strongly disagree” |
|  | People in neighborhood do not share the same values | Strongly agree; Agree; Neither agree nor disagree; Disagree; Strongly disagree | 1= “Strongly agree”; 2= “Agree”; 3=_ “Neither agree nor _disagree”; 4=_“Disagree”; 5= “Strongly disagree” |
| **NEIGHBORHOOD AND PHYSICAL ENVIRONMENT** |  |  | From the aggregate sum of the following 8 items, divided into tertiles: 0 = tertile 1-2; 1= tertile 3 |
|  | Is there an exercise facility in your neighborhood? | Yes; No | 0 = "No"; 1 = "Yes" |
|  | Is there a park in your neighborhood? | Yes; No | 0 = "No"; 1 = "Yes" |
|  | Is there a grocery store in your neighborhood? | Yes; No | 0 = "No"; 1 = "Yes" |
|  | Is there a fast-food restaurant in your neighborhood? | Yes; No | 0 = "No"; 1 = "Yes" |
|  | Is there a sit-down restaurant in your neighborhood? | Yes; No | 0 = "No"; 1 = "Yes" |
|  | Is there a subway, bus, or trolley stop in your neighborhood? | Yes; No | 0 = "No"; 1 = "Yes" |
|  | Are there sidewalks in your neighborhood? | Yes; No | 0 = "No"; 1 = "Yes" |
|  | Are there walking/and or bicycle paths in your neighborhood? | Yes; No | 0 = "No"; 1 = "Yes" |
| **HEALTHCARE** | Do you have a usual source of healthcare? | Yes; No | 0 = "No"; 1 = "Yes" |
|  | Do you have health insurance? | Yes; No | 0 = "No"; 1 = "Yes" |
|  | Overall, how hard has it been for you to get the health services you have needed? | Very hard; Fairly hard; Not too hard; Not hard at all | 0= “Not too hard”; “Not hard at all”  1= “Very hard”; “Fairly hard” |
