## Supplementary Table 2 for "The Association Between Social Determinants of Health and Alzheimer Disease Blood Biomarkers in Midlife"

**Supplementary Table 2.** Quality control (QC) results for plasma p-tau217, Aβ1-42, and Aβ1-40 across 15 analytical runs.

| **QC Sample** | **p-tau217 (pg/mL), Mean** | **%CV** | **Aβ1-42 (pg/mL), Mean** | **%CV** | **Aβ1-40 (pg/mL), Mean** | **%CV** |
| --- | --- | --- | --- | --- | --- | --- |
| Con 1 | 0.452 | 3.76 | 20.34 | 3.84 | 212.88 | 2.87 |
| Con 2 | 3.638 | 4.48 | 199.87 | 3.14 | 2243.72 | 1.67 |
| Pool 3 | 0.212 | 10.15 | 13.94 | 8.93 | 154.73 | 4.69 |
| Pool 4 | 0.521 | 5.21 | 19.14 | 6.29 | 266.26 | 3.34 |
| Pool 5 | 0.118 | 9.49 | 22.48 | 5.53 | 244.47 | 3.31 |
| Pool 6 | 0.400 | 7.01 | 19.66 | 7.05 | 236.02 | 2.58 |

Abbreviations: %CV, coefficient of variation; QC, quality control.

Con 1 and Con 2 represent manufacturer-provided aqueous controls at low and high concentration ranges. Pools 3–6 are plasma-based QC samples spanning normal and abnormal biomarker profiles. Mean concentrations (pg/mL) and coefficients of variation (%CV) are reported, demonstrating consistent assay performance across all analytes.
